# Effects of different cluster-set rest intervals during plyometric-jump training on measures of physical fitness: a randomized trial

**DOI:** 10.1101/2023.04.16.23288651

**Authors:** Behzad Taaty Moghadam, Hossein Shirvani, Rodrigo Ramirez-Campillo, Eduardo Báez-San Martín, Ali Abdolmohamadi, Behzad Bazgir

**Affiliations:** Exercise Physiology Research Center, Lifestyle Institute, Baqiyatallah University of Medical Sciences, Tehran, Iran; Exercise and Rehabilitation Sciences Institute. School of Physical Therapy. Faculty of Rehabilitation Sciences. Universidad Andres Bello. Santiago, postal code 7591538, Chile; Sports Coach Career, School of Education, Universidad Viña del Mar, Viña del Mar 2520000, Chile; Department of Sport Sciences, Faculty of Physical Activity and Sport Sciences, Universidad de Playa Ancha, Valparaíso 2340000, Chile; University of non-governmental Qadir, Langrod, Iran

**Keywords:** Stretch-shortening cycle exercise, Physical functional performance, resistance training, muscle fatigue, body composition

## Abstract

The optimal intra-set rest in cluster sets (CLS) plyometric-jump training (PJT) to improve physical fitness remains unclear. Thus, this study compared the effects of PJT with traditional (TRS) vs. CLS structures using different intra-set rests on physical fitness components. Forty-seven recreationally active young men performed 3-5 sets of 10-12 repetitions of upper- and lower-body exercises twice a week for six weeks using different set configurations as the TRS group (no intra-set rest), and the CLS10, CLS20 and CLS30 groups with 10, 20 and 30 s intra-set rest, respectively, while the total rest period (i.e., 180 s) was equated. Testing was carried out 48 h before and after the intervention and the rating of fatigue (ROF) was also assessed 20 min after the first and last session. There was no significant difference in the mean energy intake between groups (*p* > 0.05). The ANCOVA revealed that all groups showed similar improvements (*p* < 0.05) in body mass, body mass index, fat-free mass, one repetition maximum (dynamic strength) and repetitions to failure (muscular endurance) in back squat and chest press, handgrip strength, standing long jump, 20 m sprint, and 9-m shuttle run (change of direction speed), whereas the ROF decreases were greater in the CLS20 and CLS30 groups (*p* < 0.05). Compared to the TRS structure, six weeks of PJT with an intra-set rest of 20 s, or 30 s induced similar improvements in the measures of physical fitness and anthropometrics, with lower exercise-induced fatigue perception.

## Introduction

Plyometric-jump training (PJT) mainly includes jump exercises that allow muscles to store energy during the deceleration phase of action and release it during the acceleration phase, and may be associated with muscle-tendon forces comparable to those achieved by conventional slow-speed resistance training [1, 2]. Indeed, PJT can improve muscle strength [3-7], jumping and sprinting performance [4-9], change of direction speed (CODS) [8, 10-12], and anthropometrics [13-15].

When designing a PJT program, one of the most important aspects that has received little attention is the ability to modify the structure of individual sets through manipulating the number of repetitions, training load, and rest periods contained within a set [16].Indeed, the addition of a short rest period within a set has been proposed, as cluster sets (CLS) configuration, to ensure that athletes can perform at the maximum level with lower accumulated fatigue during training sessions [16, 17].

The literature is usually focused on comparison between CLS versus the traditional sets (TRS) method regarding physical fitness. For example, Asadi and Ramírez-Campillo (2016) concluded that the CLS method was superior in jump and CODS adaptations in physically active males after six weeks of PJT [18]. In a more recent study, Yilmaz et al. (2021) showed that both CLS and TRS plyometric training enhanced sprint time and jump performance in male soccer players [19]. Despite the support for the CLS modality in improving physical fitness, the optimal intra-set rest period to improve specific physical fitness and anthropometric components is unknown. Indeed, the CLS configuration with a 10 s [19] or 30 s [18] recovery interval within a set resulted in significant improvements in performance, with no difference compared with the TRS method. Therefore, the purpose of this study was to compare the effects of PJT using the CLS method with different intra-set rest intervals on anthropometrics and physical fitness. We hypothesized that compared to shorter intra-set rest intervals, longer intra-set rest intervals would induce similar improvements in physical fitness and anthropometric factors, while inducing less muscle fatigue.

## Materials and methods

### Participants

This study supposed to be conducted on military forces, but due to the lack of access to a sufficient number of the relevant participants, the population was changed to active young men. However, no other changes were made in the inclusion and exclusion criteria for selecting the subjects. Therefore, a total of 52 recreationally active men aged 20-35 were recruited from five gyms of Rasht who met our inclusion criteria of exercising ≥2 sessions per week for the past 3 months and were not injured. None of them had any background in regular PJT or competitive sports with any kind of jumping exercises during the trial. During the study, five subjects withdrew due to personal reasons and consequently data from 47 subjects (mean ± standard deviation [SD]: age, 26.5 ± 3.9 years; body mass, 78 ± 9.2 kg; body height, 173.7 ± 7 cm; body mass index [BMI], 25.8 ± 2 kg/m^-2^; body fat, 18.8 ± 5.2%) were included in the final analysis (Fig 1). Using a random-numbers table and a simple type of randomization, subjects were randomly divided into four groups including TRS (no intra-set rest, n = 12), CLS10 (10 s intra-set rest, n = 11), CLS20 (20 s intra-set rest, n = 13), and CLS30 (30 s intra-set rest, n = 11) and their order to perform the measurements was random. After data collection, a random identification number was assigned to each subject by a person who was not involved in the study procedures, so that the authors had no access to information that could identify individual subjects. The present research was conducted in accordance with the Declaration of Helsinki. All subjects signed written informed consent and were made aware of the risks associated. This study was approved by the Research Ethics committees of Baqiyatallah Hospital (<u>IR.BMSU.BAQ.REC.1400.043</u>) and was registered by the Thai Clinical Trials Registry (https://www.thaiclinicaltrials.org/show/TCTR20220210007).

**Fig 1.**
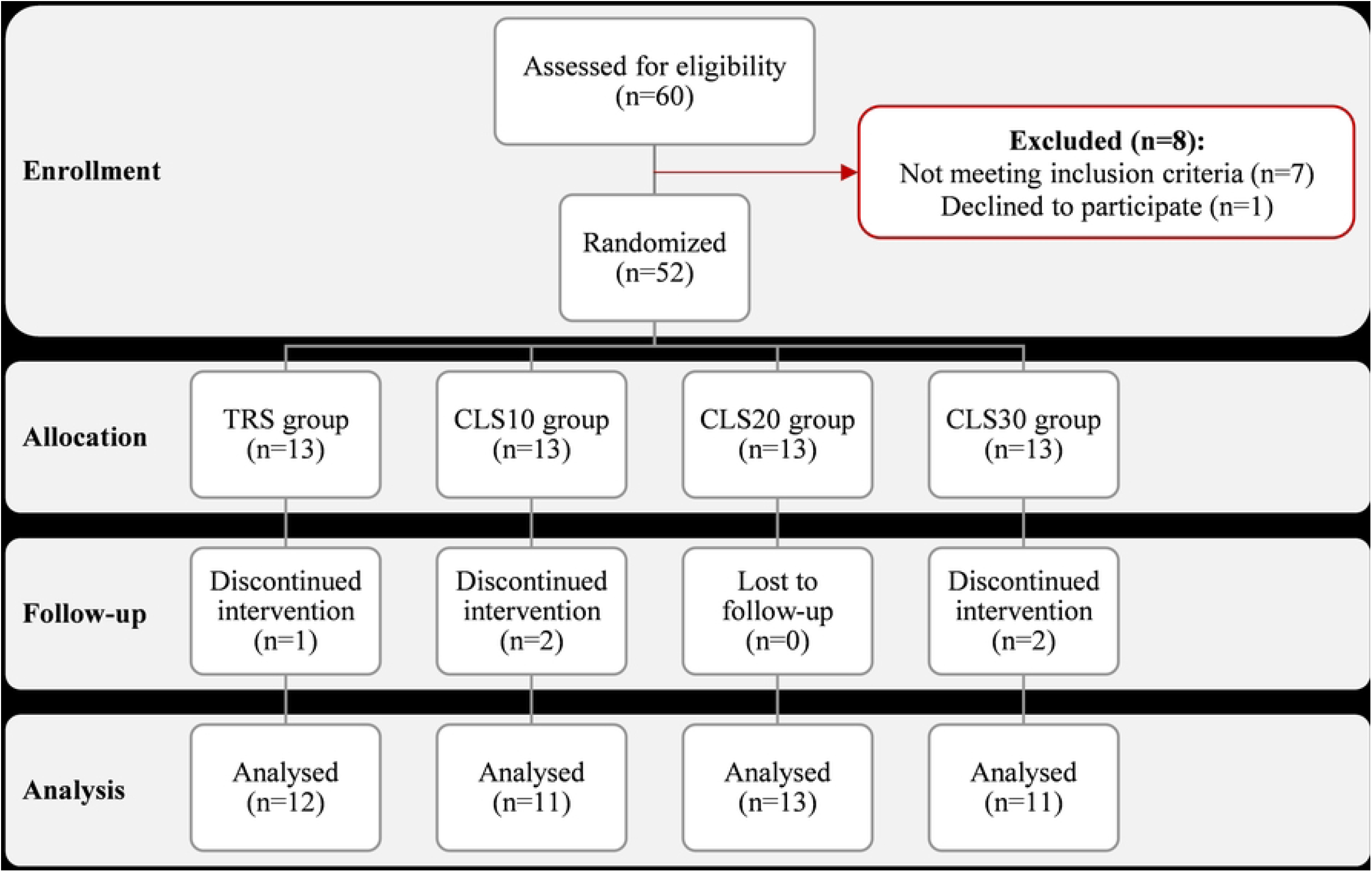
Recruitment flowchart from enrolment to data analysis.

### Procedures

This study used a between-subjects randomized design to examine the effects of PJT using the CLS structure with different rest intervals on physical fitness. The rest intervals including 0 (i.e., the TRS configuration), 10, 20, and 30 s were compared using four independent groups while other exercise prescription components were similar between them. In this model, the intervals were manipulated so that total rest time was the same for all the four groups.

### Training program

The program was performed from April 10, 2022, to May 18, 2022, every Sundays and every Wednesdays morning from 9:00-11:00. During the study period, subjects did not do any other exercise except the PJT protocol. Before the beginning of training and testing, all subjects participated in a familiarization session covering all training and testing requirements. The training protocol comprised 2 sessions per week of 4 exercises including countermovement jump, push-up jump, lateral skater jump, and incline push-up jump, across 6 weeks of training (Table 1). This training duration is proposed to ensure neuromuscular adaptations without excessive fatigue or strain [12, 18]. All training sessions were completed in a gymnasium with rubber mat surface and subjects were required to jump for maximal effort through minimizing their contact with the ground. Given that subjects did not have any history of formal plyometrics, all training sessions were closely supervised and particular attention was paid to demonstration and execution. Subjects performed a standardized warm-up at the beginning of each training session and were asked to refrain from any additional PJT or strength training throughout the study. All measurements were carried out 48 h before and after the training program at the same time of day.

**Table 1.**
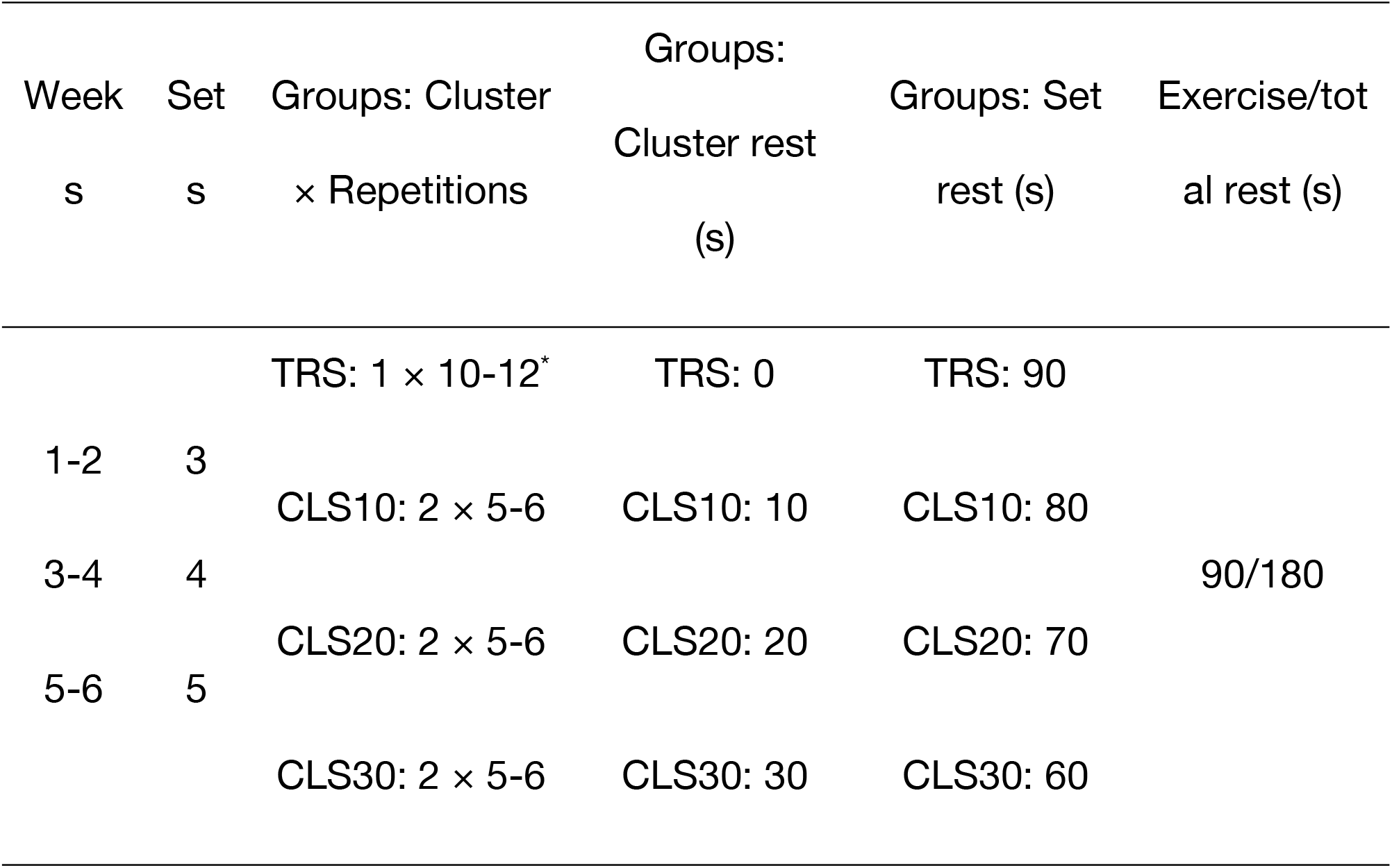
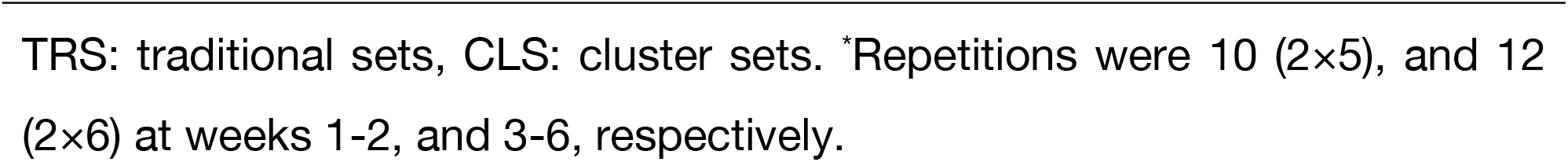
Plyometric training protocol.

| Week<br>s | Set<br>s | Groups: Cluster<br>× Repetitions | Groups:<br>Cluster rest<br>(s) | Groups: Set<br>rest (s) | Exercise/tot<br>al rest (s) |
| --- | --- | --- | --- | --- | --- |
|  |  | TRS: 1 × 10-12* | TRS: 0 | TRS: 90 |  |
| 1-2 | 3 | CLS10: 2 × 5-6 | CLS10: 10 | CLS10: 80 |  |
| 3-4 | 4 |  |  |  | 90/180 |
|  |  | CLS20: 2 × 5-6 | CLS20: 20 | CLS20: 70 |  |
| 5-6 | 5 |  |  |  |  |
|  |  | CLS30: 2 × 5-6 | CLS30: 30 | CLS30: 60 |  |

TRS: traditional sets, CLS: cluster sets. \*Repetitions were 10 (2×5), and 12 (2×6) at weeks 1-2, and 3-6, respectively.

### Anthropometric measurements

Standing height was measured to the nearest 0.5 cm. Body mass and the estimation of total body fat were determined by bioelectrical impedance analysis (BF511 Monitor, Omron Healthcare, Inc. Kyoto, Japan), based on the manufacturer’s guidelines, with a standard error of 3.5% for the estimation of body fat percentage. The BMI and fat-free mass (FFM) were also calculated from the standard equations (i.e., [body mass/height^-2^] and [body mass – body fat], respectively).

### Muscle strength test

Before the commencement of testing, a 10-min warm-up consisting of light-intensity movements and stretching and two warm-up sets of exercises with no load was performed. Lower-body (i.e., back squat) and upper-body (i.e., chest press) dynamic strength was determined as the maximum weight lifted in a single and complete repetition (one repetition maximum [1RM]) in the same order as mentioned, according to the guidelines of the National Strength and Conditioning Association [20]. In brief, subjects performed a specific warm-up set with 5 repetitions carried out at ∼50% of subject’s perceived 1RM followed by 1 to 2 sets of 2–3 repetitions at a load corresponding to ∼60–80% 1RM. They then performed sets of 1 repetition of increasing load for 1RM determination. This procedure was completed within 4 to 5 attempts and a rest period of at least 3 min was allotted between each attempt to avoid fatigue [21]. Proper lifting technique was demonstrated and practiced for each of the exercises. For evaluating grip strength on both hands, subjects were asked to maximally squeeze a hand dynamometer (78010, Lafayette, Inc. USA) without ancillary body movements, while the elbow was flexed to 90° at the side of the body. Two attempts were recorded (to the nearest 1 kg) with each hand, with at least 2 min of rest between them, and the highest records were used for the final analyses. In addition, at least 5 min of passive rest separated each strength test.

### Standing long jump (SLJ)

Subjects performed the SLJ test two times, and the best record was used for data analysis. They were asked to execute a two-legged horizontal jump as far as possible out of a standing position with swinging arms and flexing knees. The distance from the beginning of the starting line to the most posterior aspect of the subject’s body was measured in cm by the same examiner.

### Change of direction speed (CODS) and linear sprinting speed tests

The CODS performance was determined by a 9-m shuttle run test on a hardcourt track marked with tapes and pylons. During the test, subjects sprint forward for 9 m and come back to the starting line. They passed the distance four times and had to touch the tapes at the end of the first three 9-m distances with either the left or the right hand. For assessing sprinting ability, a linear 20 m sprinting test was performed. In doing so, a standard hardcourt sprint track of 20 m length was prepared by setting up pylons and the starting and finishing lines were marked by white tapes on the floor. At the end of both tracks, there was enough space for subjects to continue moving until they stopped completely. Subjects started out of a standing position and had two attempts for each test with a 2-3 min rest period. A standard stopwatch was used to record time and the best time in seconds was used for data analysis. All measurements were conducted by the same examiner (blinded regarding subject’s groups allocation) who had practiced many times to become fully familiar with the device and to reduce the possible error of measurements.

### Local muscular endurance test

Subjects performed a set of maximum number of repetitions at 60% of 1RM. Once the back squat testing was finished, the chest press was conducted after a 4-5 min passive rest [21, 22]. Subjects were encouraged to perform as many repetitions as possible using the proper lifting techniques and the total number of proper repetitions performed was recorded.

### Perceived fatigue

This was determined using a new 11-point numerical scale ranging from zero to ten, called rating of fatigue (ROF), effective and valid in assessing changes in fatigue in a variety of contexts [23]. Twenty minutes after the first and last training session, each subject rated how fatigued they felt according to the numbers from 0 (referred to not fatigued at all) to 10 (referred to total fatigue and exhaustion). All subjects were thoroughly explained how to rate their perceived fatigue during the familiarization session.

### Diet control

Subjects were asked to maintain their habitual diets throughout the study. Written and verbal instructions were provided so that subjects could record the type and portion sizes of daily foods consumed 48 h before pre-test measurements. They also were instructed to mimic this diet 48 h before the post-test measurements. Dietary data analysis revealed no significant differences between groups before pre- and post-test sessions (Fig 2).

**Fig 2.**
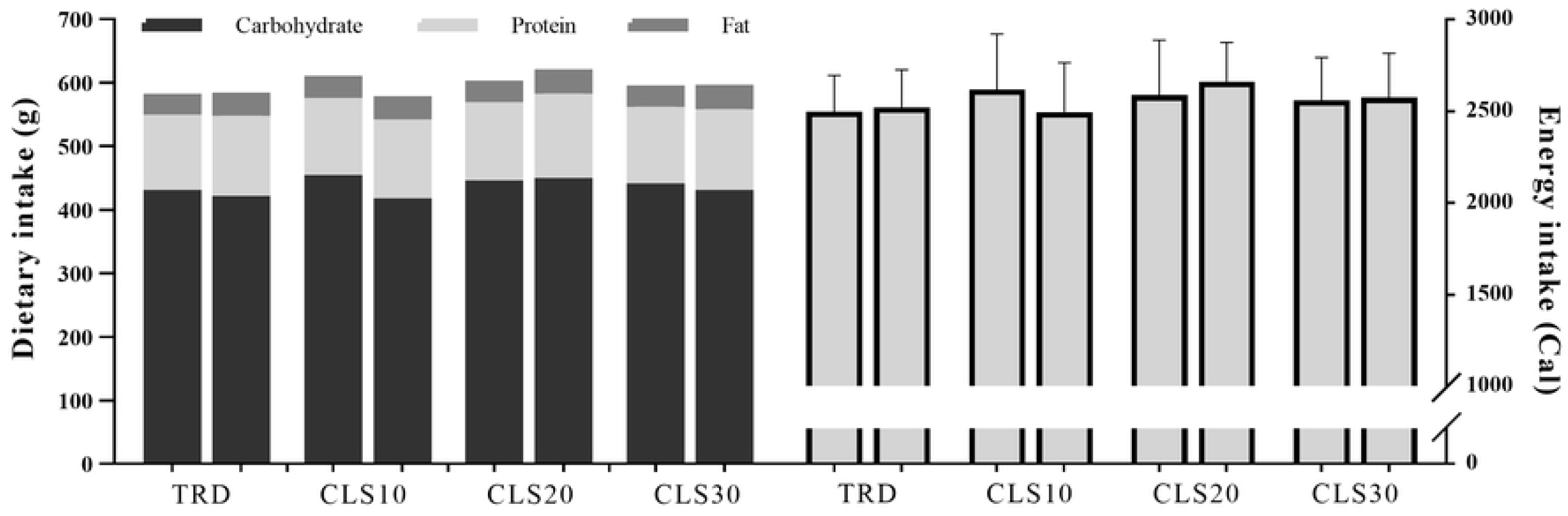
Mean of dietary and energy intake 48 h before pre-testing (left column for each group) and post-testing (right column for each group).

### Statistical analysis

The sample size was calculated using an a priori power analysis by G*Power software, version 3.1.9.4. Given a statistical power (1-ß error) of 0.9 and a moderate effect size (ES) of 0.6, the total sample size resulted in 44 subjects. However, considering a 20% drop out rate, the minimal sample size was set at 52 subjects. The Statistical Package for Social Science (SPSS, v. 22^®^, Inc. Chicago, IL) was used to analyze data at an alpha level of *p* ≤ 0.05 for all tests. Pre- and post-intervention values for each dependent variable were analyzed to determine if the distributions were normal using the Shapiro-Wilk test. A paired-samples *t* test was applied to establish if there was a potential difference in each group’s mean values from pre- to post-training (within-group analysis). An analysis of covariance (ANCOVA) was conducted for the dependent variable difference scores (post – pre) with pre-test values as covariates. Moreover, Cohen’s *d* was also calculated as ES statistic and the magnitudes were considered trivial (< 0.2), small (0.2 ≤ *d* < 0.5), medium (0.5 ≤ *d* < 0.8), and large (0.8 ≤) [21].

## Results

Means and SDs for body mass, BMI, body fat percentage, and FFM are listed in Table 2. The within-group comparisons indicated a significant reduction in body mass and BMI and an increase in FFM for all groups following the training program. However, no significant differences were found between groups in any of the outcomes (*p* > 0.05).

**Table 2.**
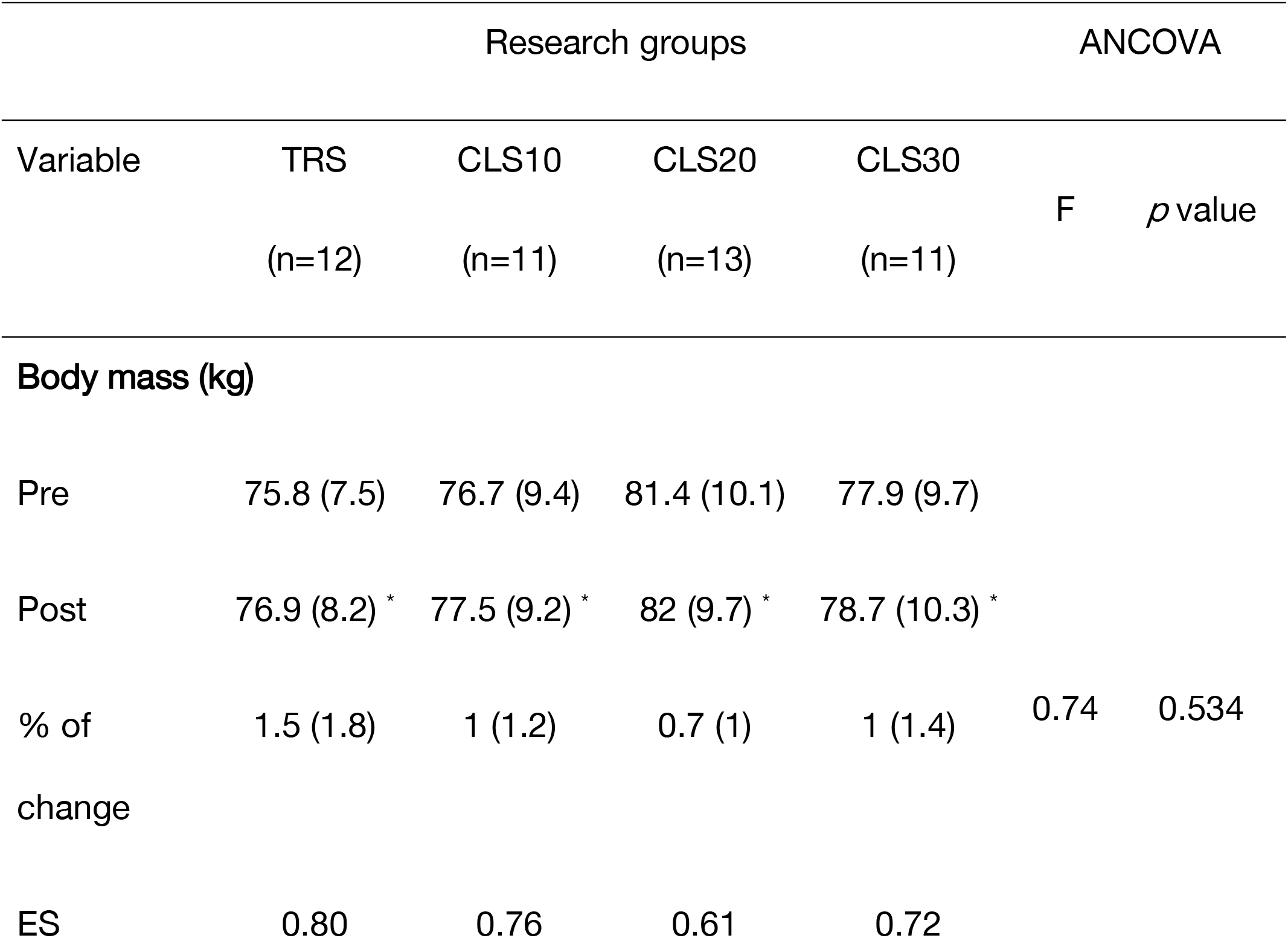
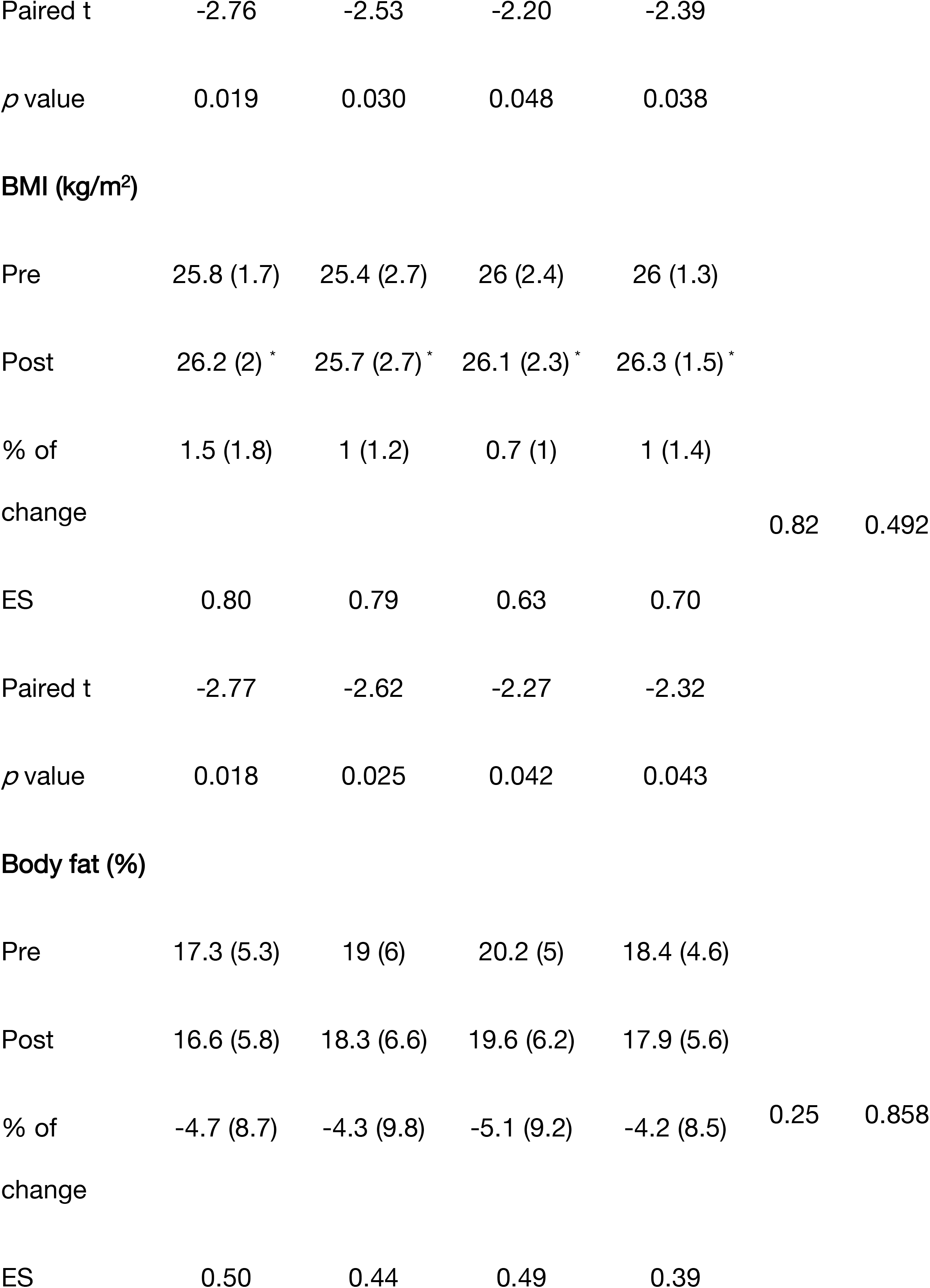
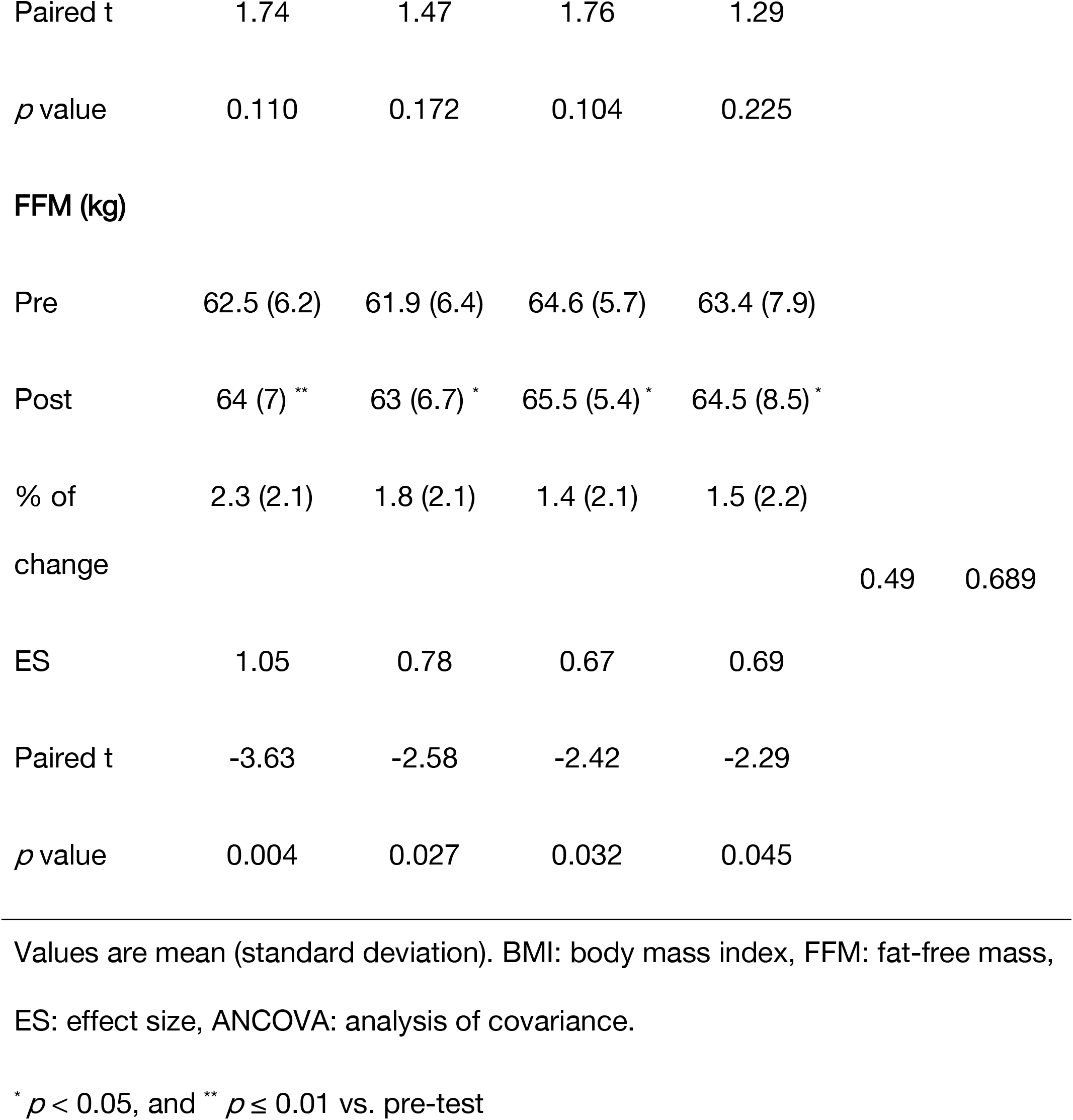
Changes in anthropometric outcomes following six weeks of plyometric training in traditional sets (TRS) group with no intra-set rest, and three cluster sets (CLS) groups with 10 (CLS10), 20 (CLS20), and 30 (CLS30) seconds intra-set rest.

Compared to their pre-training values, all four training groups showed significant improvements in relative lower- and upper-body strength, relative handgrip strength, local muscular endurance, SLJ, linear sprint, and the CODS performance, with no difference between groups (Fig 3-a to 3-j). Although ROF scores were significantly decreased (improved) from pre- to post-training in all four training groups, the CLS20 and CLS30 groups exhibited greater improvements compared to the TRS and CLS10 groups (Fig 4).

**Fig 3.**
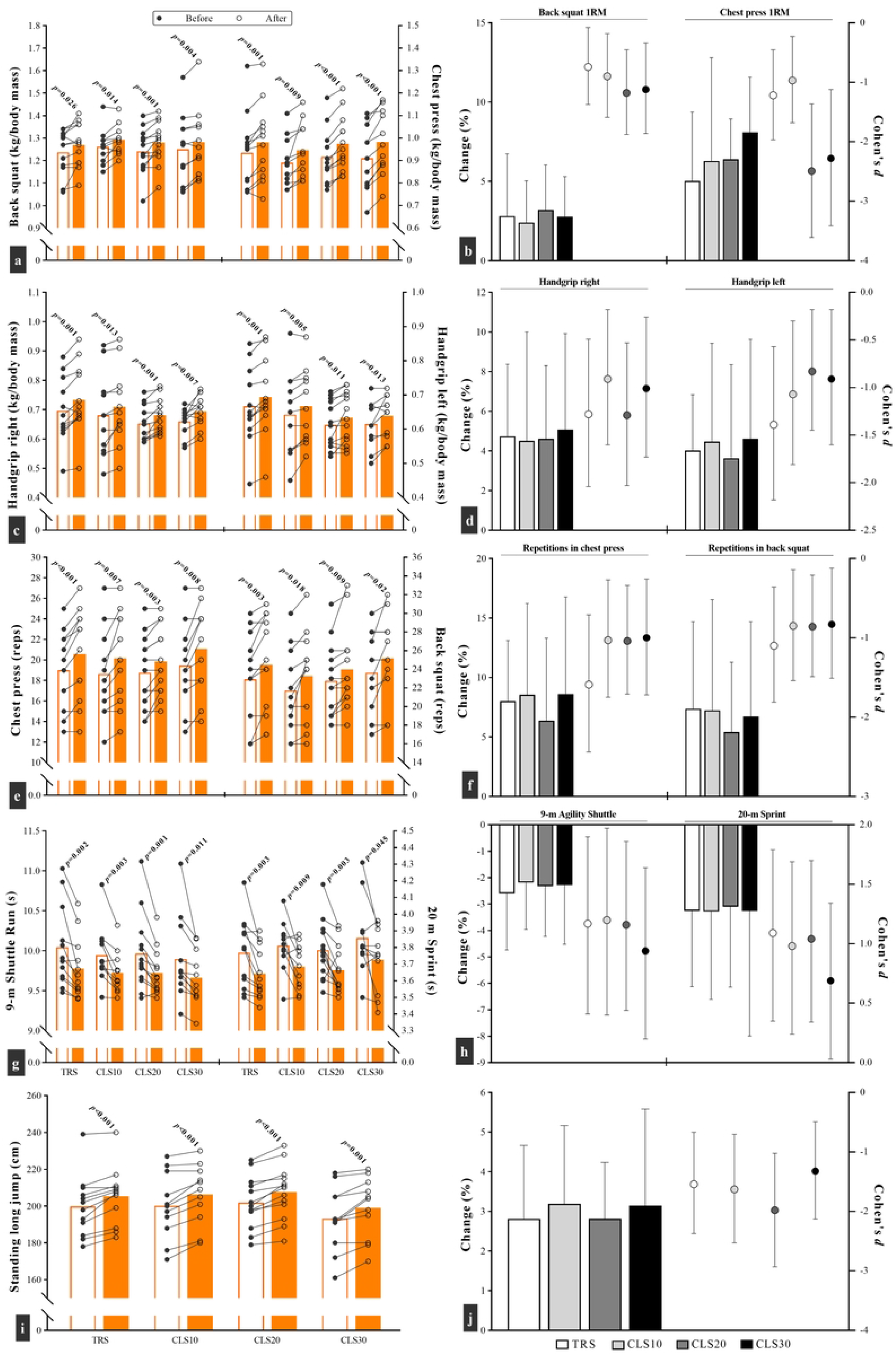
Changes in performance parameters, and percentage change and effect size for a-b) muscle strength, c-d) handgrip strength, e-f) local muscular endurance, g-h) sprint and change of direction speed, and i-j) standing long jump, after six weeks of plyometric jump training in traditional sets (TRS) group with no intra-set rest, and three cluster sets (CLS) groups with 10 (CLS10), 20 (CLS20), and 30 (CLS30) seconds intra-set rest. Fig a, c, e, g, i: white and orange columns denote outcomes results before and after the intervention. Moreover, each circle corresponds to a single subject. Fig b, d, f, h, j: the columns and circles correspond to the left and right y axis, respectively.

**Fig 4.**
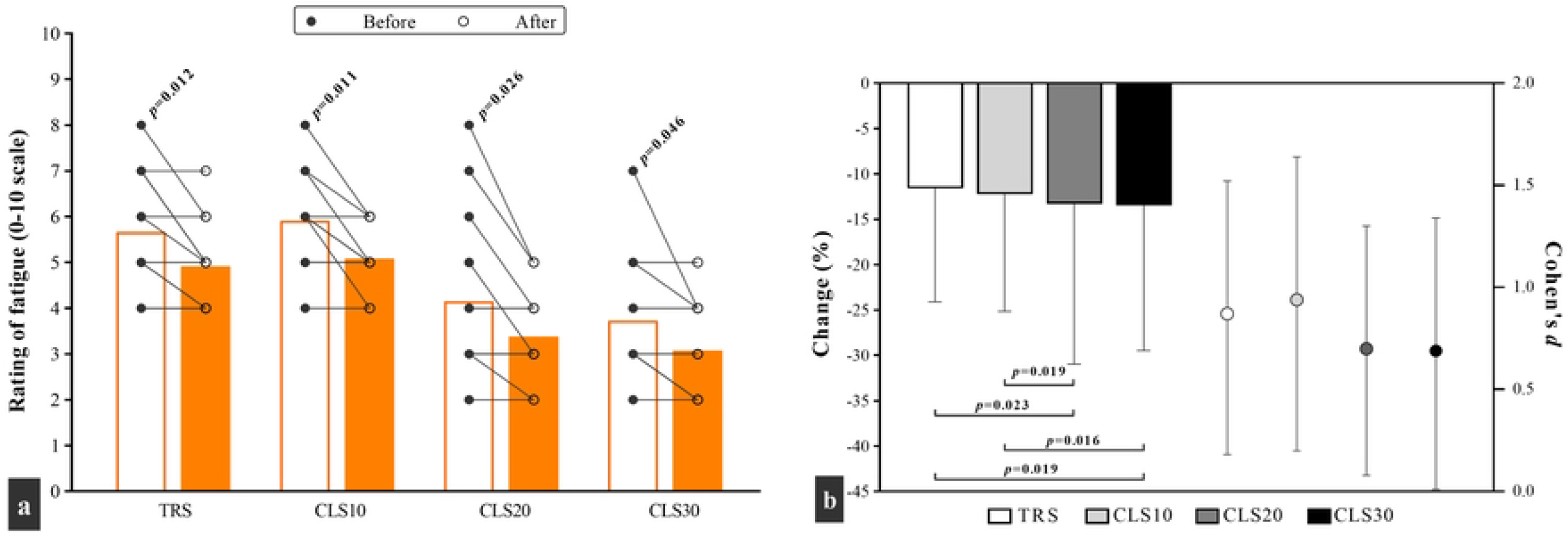
Changes in a) the rating of fatigue, and b) percentage change and effect size, after six weeks of plyometric jump training in traditional sets (TRS) group with no intra-set rest, and three cluster sets (CLS) groups with 10 (CLS10), 20 (CLS20), and 30 (CLS30) seconds intra-set rest. Fig a: white and orange columns denote outcomes results before and after the intervention. Moreover, each circle corresponds to a single subject.

## Discussion

In recent years, PJT protocols have been modified by incorporating a brief intra-set rest period (i.e., 10 to 30 s) aimed toward lower exercise exertion. Such protocols, called CLS configuration, have been shown to be effective in enhancing sprint, jump and the CODS performance [18, 19]. In the present study, we compared the effects of different rest intervals during the CLS structure used in PJT on physical fitness factors in recreationally active men. In fact, to our knowledge, this is the first study that has investigated physical fitness adaptations to manipulating the intra-set rest period.

After 6 weeks, we observed that body composition variables changed in all groups, except body fat percentage. Although there was no statistically significant difference between groups, the TRS group showed large ESs body mass and BMI reduction and FFM increase compared to medium ESs for the CLS groups. These findings suggest that short-term PJT without intra-set rest recovery may be more effective in improving body composition through increasing FFM, with no effect on body fat percentage. These observations agree with previous findings, indicating that PJT is associated with increasing FFM without changing fat mass [24]. In contrast, MacDonald et al. (2012) reported a significant gain in body fat percentage following 9 weeks of TRS plyometric training, which these changes were probably attributed to changes in subjects’ diet rather than the training protocol [14]. In another study [25], PJT did not change BMI in volleyball players that is in contrary to the present findings. This may be attributed to differences in training program (upper-body vs. upper- and lower-body plyometrics in our study) and subjects (professional volleyball players vs. recreationally active men in our study). Professional athletes achieve adaptations in which further alterations in body composition may require higher intensity and volume of training.

Fig 3-a to 3-d represent the changes in relative strength measures that occurred in groups after 6 weeks of training. The ES statistic obtained for the CLS20 (back squat, *d* = -1.2; chest press, *d* = -2.5) and CLS30 (back squat, *d* = -1.1; chest press, *d* = -2.3) groups was greater than the TRS (back squat, *d* = -0.74; chest press, *d* = -1.2) and CLS10 (back squat, *d* = -0.9; chest press, *d* = -1) groups. Nonetheless, similar results were not observed for the right- and left-hand handgrip. These results were further supported by previous studies indicating the effectiveness of plyometrics in enhancing maximal dynamic strength [14, 25, 26]. Although it has been proposed that 6 weeks of PJT may not be enough to induce hypertrophic changes [14], our findings show a significant 1.4 to 2.3% increase in FFM that can almost be indicative of hypertrophic effects. Thus, in addition to neural adaptations, it can be conceived that part of the increase in muscle strength observed in this study may be due to an increase in muscle mass. Altogether, further research with a longer duration is needed to establish hypertrophy-induced increase in muscle strength after PJT.

A novel finding in the present study was that all groups significantly improved local muscular endurance in chest press (6.4-8.6%) and back squat (5.4-7.4%), whereas the respective ES values were high, with a greater magnitude for the TRS group than others (Fig 3-e and 3-f). These results are important due to the lack of sufficient data in this area. Indeed, despite contributing to muscle strength, power and sprint as was previously confirmed [2, 14, 18, 19], we here found that PJT using both TRS and CLS configurations was able to enhance maximum number of repetitions to failure at 60% of 1RM, which in line with the study conducted by Al Ameer (2020) even though the muscle group examined in that study was different. The author reported that a 12-week twice weekly PJT program resulted in better sit-ups records in adult soccer players compared with pre-tests [27]. An important point to note is that we re-adjusted the load in the post-study assessment according to the newly established 1RM, which can partially eliminate the effects of increasing muscle strength on the results. Therefore, the improved local muscular endurance in this research would likely be related to an enhanced oxidative and buffering capacity, an increase in capillarization and mitochondrial volume, and an improve in metabolic enzyme activity [28].

All groups significantly enhanced the CODS (2.2-2.6%) and linear sprint (3.1-3.3%) performance, but the respective ESs were moderate to high (Fig 3-g and 3-h). Although some authors have reported no effect of PJT on 20 m sprint [29] and the CODS [30, 31] records, our findings add consistently to previous works showing that sprint ability and the CODS performance improved by 2.7% and 3.8% [19], and 6.3% and 5.5% [18], respectively, following 6 weeks of PJT with the CLS structure. These improvements might be attributed to several reasons. First, the increased muscle strength of the study subjects might offer one explanation. Specifically, the increased back squat strength occurred in this study can positively transferred into sprint ability [32] and is also strongly correlated with CODS [33]. The second explanation is related to neuromuscular adaptations such as greater motor unit recruitment which in turn enhance muscle power output and force development ability [12, 18]. Third, PJT may induce positive effects on key elements of sprint and the CODS tasks including lower ground contact times as a result of high muscular force output [18, 34], the rate of force development, and the increased efficiency in using the stretch-shortening cycle (SSC) during ballistic tasks [8] even though we did not measure these parameters.

Another important change observed in this study was the increase in the SLJ distance (2.8-3.2%) in all four groups after the training period (Fig 3-i). The calculated ES statistics ranged between 1.3 and 2, while the greater magnitude was related to the CLS20 group (Fig 3-j). Most previous studies have found significant improvements in jumping performance as a result of short-term PJT [4, 18, 19, 34-36], supporting our results and the principle of specificity in training. There are several explanations for the positive effects of plyometrics, mainly related to neural adaptations, including changes in muscle structure and individual fiber mechanics, enhanced neuro-muscular coordination, changes in mechanical stiffness properties of tendons and the greater efficiency to utilize the muscles SSC [18, 34]. Regardless of these reasons, part of the improvements may be due to the increase in FFM of subjects in all groups and subsequent increase in muscular power output and the rate of force development.

Regarding ROF differences between groups (Fig 4), our results reveal a significant decrease in ROF scores in all groups, but the magnitude of decreases in the CLS20 (13.3%, *d* = 0.7) and CLS30 (14.8%, *d* = 0.7) groups was significantly greater than those of TRS (11.6%, *d* = 0.87) and CLS10 (12.2%, *d* = 0.94). In fact, the ROF score in all groups was lower after training, even considering that the number of sets and repetitions were greater in the last training session compared to the first week. Importantly, despite the total rest period (i.e., 180 s) was similar among groups, subjects who performed PJT with a 20 or 30 s intra-set rest interval felt less exercise-induced fatigue. Here, we used the ROF scale that is a valid, simple and sensitive instrument capable of tracking fatigue perception through exercise and recovery [23]. Depletion of muscle creatine content [37], pH drop due to lactate accumulation, and muscle glycogen depletion [38] can be among the possible metabolic causes of exercise-induced fatigue. By continuing to jump during plyometric exercise, tension develops in muscles and they provide the necessary force to continue exercise through increasing motor unit recruitment and firing frequency [38]. Greater motor unit recruitment, in turn, can increase more signals to the sensory cortex [38, 39] and subsequently increase the perceptions of exertion and fatigue. However, it is difficult to discuss more about these metabolic and neuromotor factors because we did not assess them independently. Therefore, more research needs to be done comparing the PJT protocols in the CLS and TRS methods with respect to the mentioned variables

## Conclusions

A six-week upper- and lower-body PJT program improved physical fitness and anthropometric measures in active young men, independent from set rest configurations. However, a CLS20 and CLS30 configuration seems to achieve greater improvements in fatigue perception reduction. The present findings provide interesting practical relevance in designing optimal plyometric training protocols using the CLS or TRS configuration for optimal physical fitness and body composition improvements, in line with reduced perceived fatigue, a key element for long-term physical activity habits.

## Data Availability

All relevant data are within the manuscript and its Supporting Information file

## Supporting information

**S1 File. PLOS’ questionnaire on inclusivity in global research**. (DOCX)

**S2 File. CONSORT checklist**. (PDF)

**S3 File. The study protocol**. (PDF)

**S4 File. Excel datasets for dietary data analysis**. Fig 2 is released from these datasets. (XLSX)

**S5 File. Excel datasets for anthropometric and physical fitness factors analysis**. Table 2, and Fig 3 and 4 are released from these datasets. (XLSX)

## Acknowledgments

The authors would like to thank all subjects who volunteered to participate in this research. We also are grateful to Dr. Nader Samami for his invaluable contribution to the study.

## Author Contributions

**Conceptualization:** Behzad Taaty Moghadam, Hossein Shirvani, Behzad Bazgir.

**Data curation:** Behzad Taaty Moghadam. Ali Abdolmohamadi.

**Formal analysis:** Behzad Taaty Moghadam, Rodrigo Ramirez-Campillo, Eduardo Báez-San Martín.

**Investigation:** Behzad Taaty Moghadam, Rodrigo Ramirez-Campillo, Eduardo Báez-San Martín.

**Methodology:** Behzad Taaty Moghadam, Hossein Shirvani, Behzad Bazgir.

**Project administration:** Behzad Taaty Moghadam.

**Resources:** Hossein Shirvani, Behzad Bazgir, Rodrigo Ramirez-Campillo.

**Software:** Behzad Taaty Moghadam.

**Supervision:** Hossein Shirvani, Behzad Bazgir.

**Validation:** Behzad Taaty Moghadam, Behzad Bazgir, Rodrigo Ramirez-Campillo, Eduardo Báez-San Martín.

**Visualization:** Behzad Taaty Moghadam.

**Writing – original draft:** Behzad Taaty Moghadam, Ali Abdolmohamadi, Behzad Bazgir.

**Writing – review & editing:** Rodrigo Ramirez-Campillo, Eduardo Báez-San Martín.

## Notes

### Competing Interest Statement

The authors have declared no competing interest.

### Clinical Trial

TCTR20220210007

### Author Declarations

The present research was conducted in accordance with the Declaration of Helsinki. All subjects signed written informed consent and were made aware of the risks associated. This study was approved by the Research Ethics committees of Baqiyatallah Hospital (IR.BMSU.BAQ.REC.1400.043) and was registered by the Thai Clinical Trials Registry (https://www.thaiclinicaltrials.org/show/TCTR20220210007).

